# Cross-Border Vaccine Supply to Conflict-Affected Darfur: A Humanitarian Lifeline through Chad - An Implementation Case Study

**DOI:** 10.64898/2026.04.01.26349918

**Authors:** Victor Sule, Dalya Eltayeb, Hashim Eltayeb, Khattab Obaid, Ismael Alshekh, Mohammed Alhaboub, Abdulaziz Abdelrahman Adam, Tedbabe Degefie Hailegebriel

## Abstract

Protracted conflict in Sudan since April 2023 has severely disrupted routine immunization services, particularly in the Darfur region, resulting in widespread vaccine stockouts, declining coverage, and increased risk of vaccine-preventable disease outbreaks. Traditional national supply routes became largely inaccessible, exacerbating inequities in immunization access for conflict-affected and displaced populations.

This paper examines the design, implementation, and outcomes of a cross-border vaccine deployment strategy implemented in 2025 through Chad to restore vaccine availability in Darfur. Using programmatic data, shipment records, coverage reports, and partner monitoring outputs, the study assessed the operational feasibility, partnership arrangements, and public health impact of the intervention on routine immunization and outbreak response.

In 2025, nearly 20 million doses of vaccines were successfully delivered to the five Darfur states through cross-border operations, supporting routine immunization services and outbreak response campaigns. Average coverage for the first dose of a DPT-containing vaccine (DPT1) increased from 22.6% in 2024 to 83.2% in 2025, while DPT3 and MCV1 coverage rose to 55.4% and 50.4%, respectively. Oral cholera vaccine campaigns achieved 90.4% coverage among targeted populations, and polio outbreak response campaigns exceeded 100% administrative coverage, reflecting both successful reach and uncertainties in target population estimates due to population displacement. Investments in cold chain infrastructure and strengthened coordination among government, UNICEF, Gavi, and implementing partners were critical to these outcomes.

The findings demonstrate that cross-border vaccine deployment can serve as a viable and effective mechanism for restoring immunization availability and support recovery of immunization service delivery in a highly constrained conflict setting. While not a substitute for functional national systems, such approaches are essential life-saving interventions during acute crises and should be integrated into preparedness planning for fragile and conflict-affected contexts.

## Introduction and Background

Immunization is widely recognized as one of the most effective and cost-efficient public health interventions and a cornerstone of primary health care (PHC). Since the establishment of the Expanded Program on Immunization (EPI) in the 1970s, vaccines have significantly reduced childhood morbidity and mortality worldwide. In many countries with sustained high immunization coverage, diseases that once accounted for a large proportion of child deaths have been substantially reduced or eliminated. Beyond preventing vaccine-preventable diseases, immunization programs also serve as an entry point for delivering other essential maternal and child health interventions, including vitamin A supplementation, growth monitoring, and community health engagement. In countries with high vaccine Programme coverage, many of the diseases that were previously responsible for most childhood deaths have essentially disappeared [1, 2].

The effectiveness of immunization programs depends heavily on the reliability of vaccine supply chains. Vaccines must be accurately forecasted, procured, transported, stored, and distributed under strict temperature-controlled conditions to ensure potency at the point of service delivery. Effective vaccine logistics systems therefore play a critical role in ensuring that vaccines reach communities safely and on time. Disruptions in supply chain systems can rapidly lead to vaccine stockouts at service delivery points, interrupting immunization services and increasing the risk of outbreaks of vaccine-preventable diseases [4, 5].

Sudan has faced multiple humanitarian and public health crises in recent decades. However, the armed conflict that erupted on 15 April 2023 triggered an unprecedented disruption of the country’s health system. Immunization service disruption in Darfur was driven not only by insecurity but also by vaccine unavailability due to supply chain breakdown. The cross-border intervention therefore tests the hypothesis that: Restoring vaccine availability can enable recovery of immunization services even in conflict settings According to the International Organization for Migration (IOM) Displacement Tracking Matrix (DTM), as of July 10, 2025, the ongoing military clashes in Sudan have resulted in an estimated 7,666,575 internally displaced persons (IDPs) since April 15, 2023. The largest numbers of IDPs are in South Darfur (16%), North Darfur (16%), and River Nile (8%). Over half (52%) of the IDPs are children under 18 years old. [6]

The conflict contributed to the backsliding of the immunization coverage in the country with coverage of first dose of Diphtheria, Tetanus and Pertussis (DTP1/Penta1) from 94% in 2022 to 48% in 2024 resulting in close to one million zero-dose children (966,670), 76% of whom are in conflict-affected states (including the five Darfur states, three Kordofan state, Khartoum and Aj Jazirah states). The five Darfur states contributed a total of 372,064 to the zero dose in 2024 accounting for 38.5%. The figure below shows the coverage for third dose of Diphtheria Tetanus and Pertussis (DPT3/Penta3) and first dose of Measles containing vaccine (MCV1) in the last five years [7,8]

**Figure 1:**
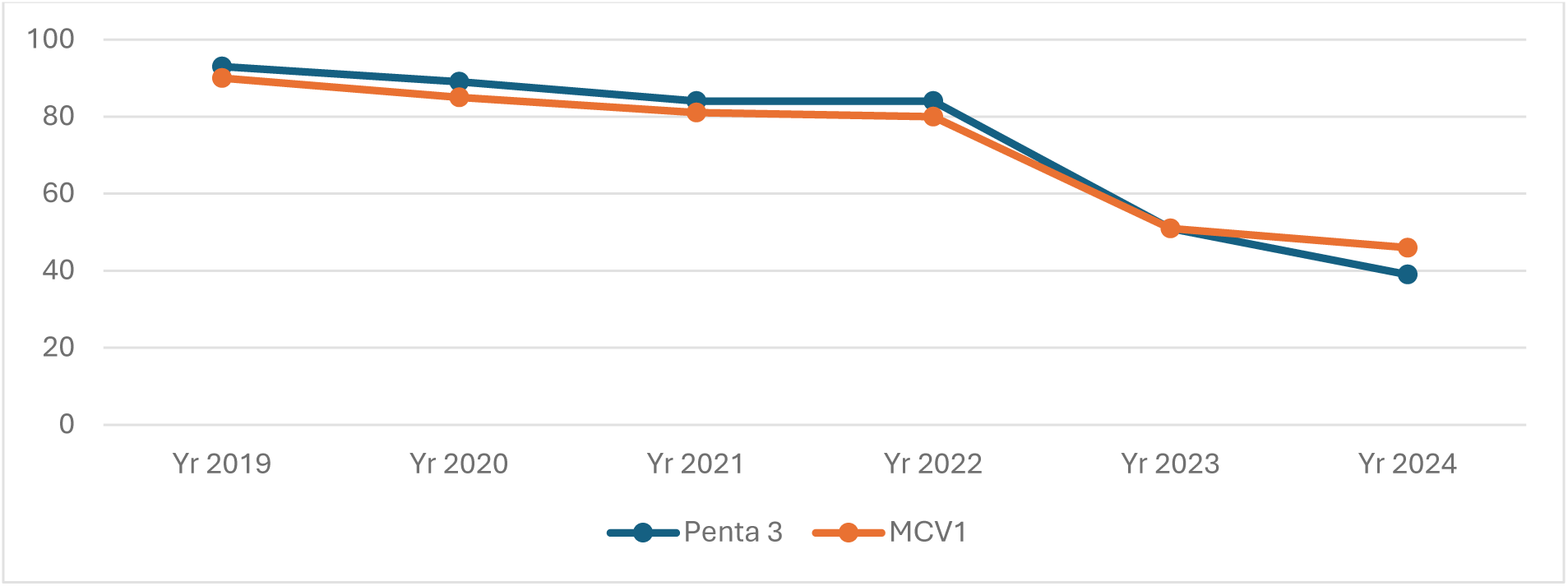
Sudan Penta 3 and MCV1 coverage 2019 -2024.

Limited vaccine availability due to conflict related access constraints contributed directly to the regression of immunization coverage in the Darfur states

Coverage in Darfur states regressed drastically contributing largely to the fall in the national routine immunization coverage, for example in South Darfur the Penta3 coverage fell to as low as 8% in 2024 from 60% in 2022. The figure below shows the coverages for Penta3 and MCV1 in the five states

**Figure 2:**
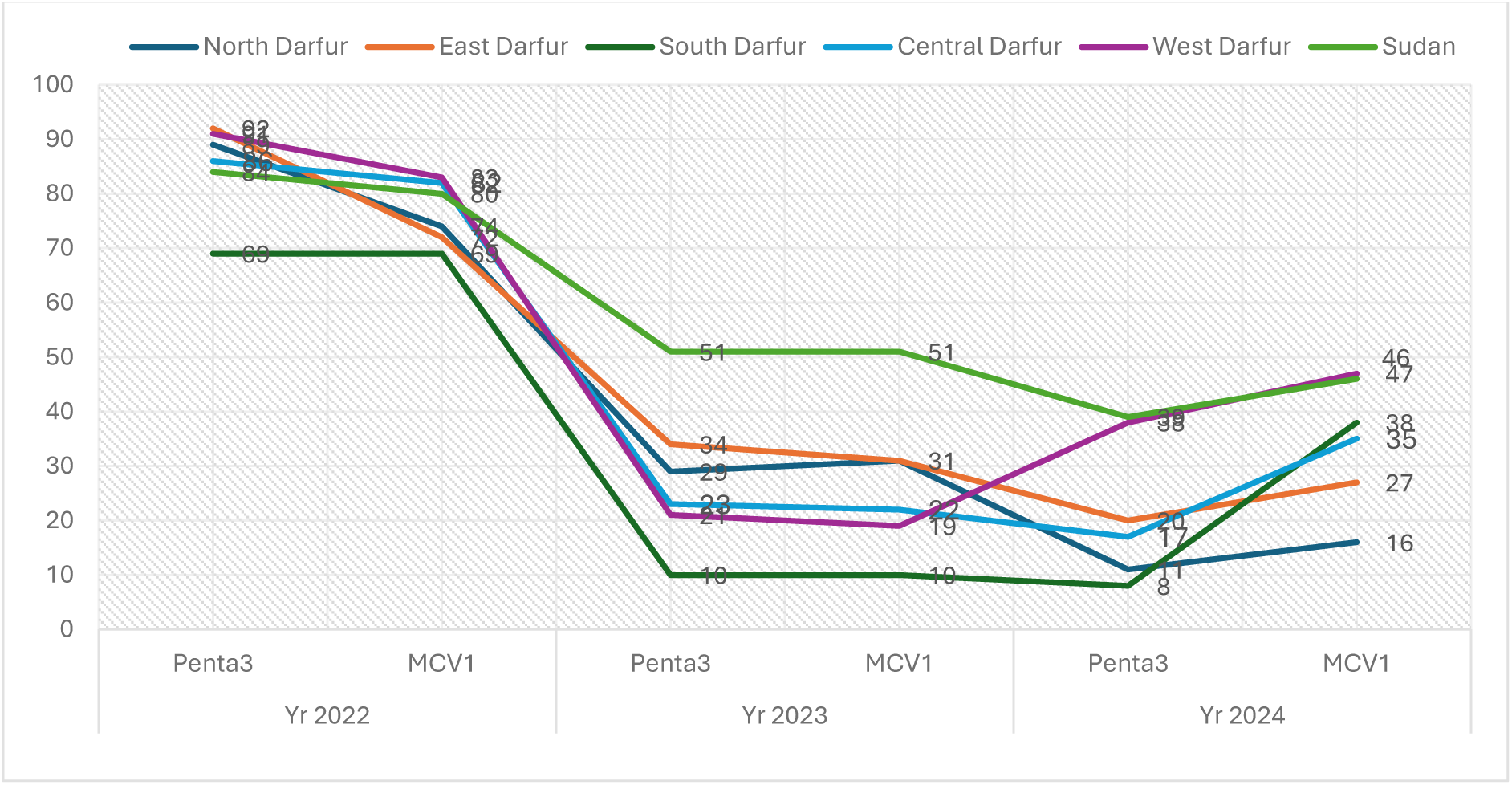
Penta 3 and MCV Coverages 2022-2024 in Darfur states [8].

The suboptimal coverage in the region has also contributed to vaccine preventable disease outbreak including measles and very recently circulating vaccine derived polio virus type 2 in two of the Darfur states pointing to the critical need to revive and sustain routine immunization services and urgent response to the outbreak to interrupt the transmission.

Since the onset of the conflict on 15 April 2023, Federal Ministry of Health (FMoH), in coordination with UNICEF undertook urgent measures to safeguard national vaccine stocks. A rapid evacuation from the central cold store in Khartoum enabled the redistribution of vaccines to 12 states excluding the five Darfur states and Khartoum effectively preventing stockouts in those regions. Darfur states were excluded from this redistribution due to recent vaccine deliveries (Quarter 2 quota) prior to the escalation of conflict and concerns about emerging insecurity in these areas. On 1 October 2023, the first shipment of vaccines to the Darfur states successfully arrived in El-Fasher, North Darfur, a result of sustained joint efforts by UNICEF and FMoH. These vaccines were used to sustain immunization services in North, West, Central, and East Darfur, while South Darfur was unable to retrieve doses due to severe insecurity at the time. Despite this breakthrough, access to Darfur remained highly constrained.[9]

From October 2023 to April 2024, UNICEF and FMoH maintained uninterrupted vaccine supply to eight states: Red Sea, Kassala, Gedaref, River Nile, Northern, Sennar, Blue Nile, and White Nile. However, conflict in Sennar in mid-2024 temporarily halted deployment to White Nile and Blue Nile, resuming only after de-escalation in December 2024. In contrast, vaccine availability in the Darfur region remained critically low, with only 8–44% of forecasted needs met in 2023, and 34–62% in 2024. South Darfur was the hardest hit, losing existing stock and remaining inaccessible for vaccine deliveries throughout 2024. East Darfur faced some disruptions due to lack of accessibility given the insecurity around major routes. West Darfur, relatively less affected, recorded the highest vaccine consumption during 2024 thanks to better access and prior stock delivery in 2024.[9]

To circumvent traditional supply bottlenecks, efforts in February 2024 focused on desert routes via Northern State. The first shipment using this path reached all Darfur states by 1 April 2024, though further deliveries were made difficult by worsening security conditions. The precarious supply chain left approximately 600,000 children under one year of age in Darfur region with 28% of Sudan population vulnerable to vaccine-preventable diseases (VPDs), with intermittent stockouts posing a serious threat of outbreaks. [9]

In December 2024, a new supply effort was launched from Port Sudan via El-Dabba in Northern State, which took some 21 days for delivery. While vaccines were successfully transported under temperature-controlled conditions, some doses showed progression with the Vaccine Vial Monitor (VVM), indicating exposure to temperature outside the recommended but still at usable stage.

Equity in immunization becomes critically strained in conflict-affected settings like Sudan, where access challenges often lead to vaccine stockouts and disrupted services. In such environments, routine immunization is compromised by insecurity, damaged infrastructure, displacement of communities, and limited humanitarian access. [10]

Children and vulnerable populations in conflict zones face a double burden: heightened risk of vaccine-preventable disease outbreaks (such as measles and polio) and reduced access to essential vaccines due to stockouts. These stockouts often result from logistical barriers, including supply chain interruptions, insecurity on transport routes, and the breakdown of cold chain systems. [10]

This prolonged disruption highlights a critical insight: immunization service decline in Darfur was driven not only by insecurity but also by vaccine unavailability resulting from supply chain breakdown. This observation suggests that restoring vaccine availability may enable recovery of immunization services even in conflict settings.

In response, alternative strategies were explored to bypass disrupted national supply routes. One such strategy involved cross-border vaccine deployment through neighboring Chad aimed at restoring vaccine availability and ensuring continued access to vaccines for vulnerable populations in the region.

Despite the growing importance of cross-border humanitarian operations in conflict settings, there is limited documented evidence on the operational feasibility and public health impact of cross-border vaccine supply mechanisms for sustaining routine immunization services. This study aims to address this gap by documenting the implementation and outcomes of a cross-border vaccine deployment strategy to Darfur.

The key objectives of the publication include:

1. Assess the impact of vaccine stockouts on immunization equity in conflict-affected border regions.
2. Document the design, implementation, and operational challenges of cross-border vaccine deployment to Darfur
3. Examine the role of partnerships among government authorities, humanitarian organizations, and international partners in enabling vaccine access across borders.
4. Generate lessons and recommendations for strengthening vaccine supply systems in fragile and conflict-affected settings.

As mentioned, this study examines the implementation and outcomes of a cross-border vaccine deployment strategy to restore vaccine availability in Darfur. The conceptual pathway below linking conflict-related supply disruptions to recovery of immunization services through cross-border deployment

**Figure 3.**
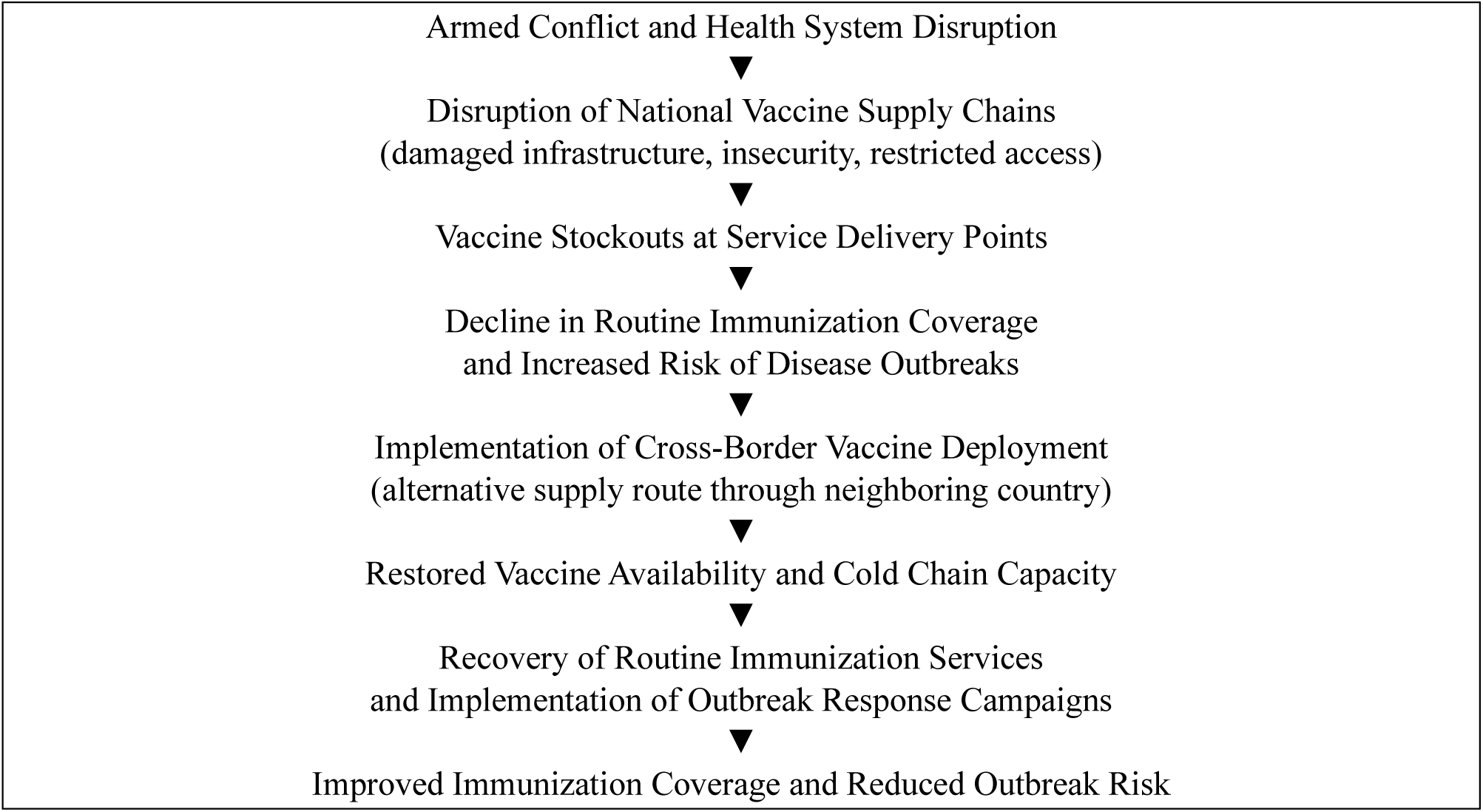
Conceptual framework illustrating how cross-border vaccine deployment can restore vaccine availability and support recovery of immunization services in conflict-affected settings.

## Method and implementation approach

### Study Design and Approach

This study presents an operational analysis of a cross-border vaccine deployment strategy implemented in 2025 to restore vaccine availability in the conflict-affected Darfur region of Sudan. The analysis draws on programmatic data, including shipment tracking records, administrative coverage reports, and partner monitoring outputs to examine the design, operationalization, and early outcomes of the intervention. This study used descriptive analysis of aggregated programmatic data; no statistical adjustment for confounding or subgroup analysis was performed

The cross-border deployment mechanism was developed in response to prolonged disruptions in national vaccine supply routes following the outbreak of armed conflict in Sudan in April 2023. Internal transport routes to the Darfur region became increasingly unreliable due to insecurity, logistical constraints, and damage to infrastructure, resulting in recurrent vaccine stockouts and interruptions in routine immunization services.

The intervention aimed to establish an alternative supply pathway through neighboring Chad in order to ensure continued vaccine availability for routine immunization and outbreak response activities in Darfur.

### Planning and Strategy Development

Planning for the cross-border vaccine deployment began in late 2024 through a series of coordination discussions between Sudan’s Federal Ministry of Health (FMoH), UNICEF, and Gavi, the Vaccine Alliance. These discussions focused on identifying feasible operational options to bypass disrupted national supply routes and restore vaccine access to the Darfur region.

Three potential operational models were considered:

1. Direct cross-border deployment: vaccines transported by air to N’Djamena International Airport in Chad and then delivered by road across the Chad–Sudan border directly to Darfur.
2. Temporary cold chain hub at the UN humanitarian hub in Abéché: vaccines stored temporarily in Abéché before onward transport into Sudan.
3. Cold chain storage at the Chad Ministry of Health facility in Abéché: installation of walk-in cold rooms to hold vaccines prior to cross-border transfer.

Following technical consultations and operational feasibility assessments, partners agreed in February 2025 to proceed with the first option—direct deployment from N’Djamena to Darfur while parallel investments were initiated to strengthen regional cold chain infrastructure in Abéché for potential future use.

### Pilot Shipment and Operational Testing

To validate the feasibility of the cross-border supply mechanism, a pilot shipment was conducted in early 2025. The pilot was designed to simulate the full operational pathway on a small scale and identify potential bottlenecks before expanding the operation. The pilot shipment assessed several key operational components, including:

- Customs clearance procedures at N’Djamena International Airport.
- Cold chain management and vaccine handover procedures between Chad and Sudan
- Security arrangements and transport logistics for overland delivery into Darfur
- Coordination mechanisms between participating organizations and national authorities

Findings from the pilot operation informed refinements to the cross-border logistics procedures and confirmed the operational feasibility of scaling up the intervention.

### Cold Chain Strengthening

Following the successful pilot shipment, additional investments were made to strengthen cold chain capacity within Darfur to support sustained vaccine deployment. UNICEF, in collaboration with the Federal Ministry of Health, relocated two 40-cubic-meter walk-in cold rooms to El Geneina in West Darfur State to serve as a regional vaccine storage hub. The facility was intended to provide buffer stock capacity and support vaccine distribution across the Darfur states.

Strengthening cold chain infrastructure was considered critical for ensuring that larger vaccine volumes delivered through cross-border shipments could be safely stored and redistributed to state and locality levels.

### Stakeholder Coordination and Roles

The cross-border vaccine deployment required coordination among multiple national and international stakeholders.

The Federal Ministry of Health (FMoH) provided national leadership and regulatory oversight, including authorization for cross-border vaccine importation and coordination of vaccine distribution within Sudan.

UNICEF, including its Sudan, Chad, and Supply Division offices, led the operational coordination of the deployment. This included vaccine procurement, international shipment arrangements, customs clearance coordination, cold chain management, and tracking of vaccine deliveries. Gavi, the Vaccine Alliance, provided financial and strategic support

At the subnational level, State Ministries of Health in Darfur were responsible for vaccine storage, distribution to localities and health facilities, while Implementation partners including Save the Children, Alight, and International Medical Corps (IMC) supported last-mile distribution, and community mobilization activities to facilitate vaccine uptake.

### Risk Assessment and Mitigation

Given the conflict context and complexity of the cross-border supply chain, several operational risks were identified during the planning phase. These included potential delays at border crossings, cold chain disruptions during transit, customs clearance delays, and coordination challenges among stakeholders.

Mitigation measures included pre-arranged permits and regulatory approvals, the use of WHO-prequalified cold chain equipment and temperature monitoring devices, designation of logistics focal points for customs clearance, and the establishment of regular coordination meetings among partners to address operational challenges in real time.

These mitigation strategies were designed to maintain cold chain integrity, ensure timely vaccine delivery, and support continuity of immunization services in the Darfur region.

### Implementation process

The conceptualization of the cross-border vaccine deployment initiative began in mid-2023 after the eruption of the conflict. Following several rounds of consultations and coordination among stakeholders, implementation of the agreed approach commenced in March 2025 with the initial test shipment that included three vaccines: BCG, Tetanus-diphtheria (Td), and Measles. to test the operational feasibility of the cross-border supply chain.

Following the pilot phase, routine cross-border shipments were initiated in June 2025. Coordination among stakeholders was maintained through weekly operational meetings involving UNICEF Sudan and Chad country offices, UNICEF Supply Division, and Gavi.

Throughout 2025, multiple shipments of routine and campaign vaccines were transported via N’Djamena and delivered overland to Darfur. These included vaccines for routine immunization as well as outbreak response, such as measles-rubella (MR), oral cholera vaccine (OCV), and novel oral polio vaccine type 2 (nOPV2).

A summary of vaccine shipments delivered through the cross-border mechanism in 2025 is presented in Table 1.

**Table 1:**
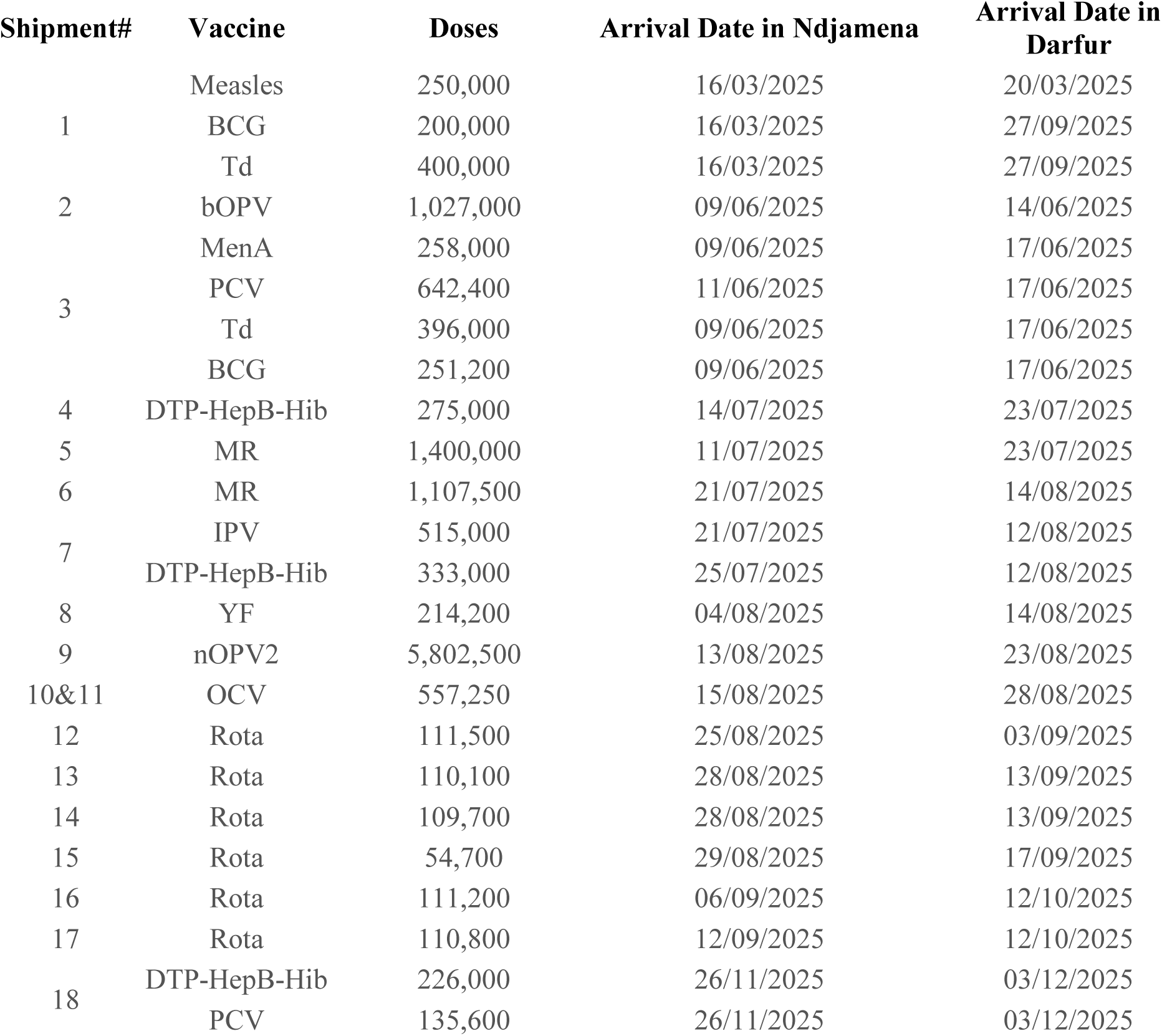
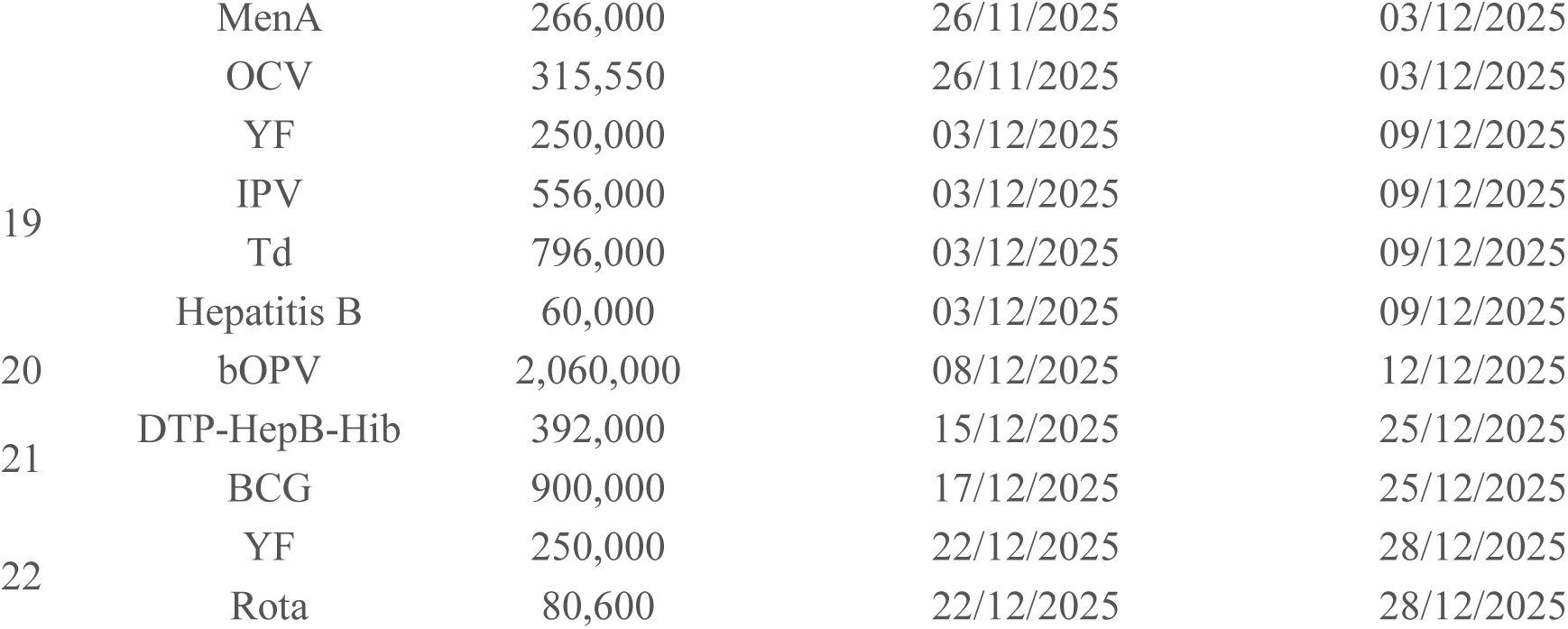
2025 Cross-Border Vaccine Shipment Summary.

### Logistics Management and Transport Arrangements

UNICEF Supply Division coordinated the international logistics for vaccine shipments, engaging a third-party freight forwarder, to manage the international transport, customs clearance, and logistical coordination of vaccine deliveries.

For onward transport from Chad into Sudan, collaboration with a local logistics provider in Chad enabled secure, temperature-controlled road transport from N’Djamena to El Geneina in West Darfur. This system ensured end-to-end cold chain integrity and compliance with regulatory requirements.

## Results

### Vaccine Supply Through Cross-Border Deployment

The implementation of the cross-border vaccine deployment mechanism enabled large-scale restoration of vaccines supply to the Darfur region during 2025. More than 17 million doses of vaccines were transported through N’Djamena International Airport in Chad and delivered to the five Darfur states to support both routine immunization services and outbreak response activities.

These deliveries represented approximately 85% of all vaccine shipments reaching the Darfur region in 2025, demonstrating the central role of the cross-border supply route in restoring vaccine availability in the conflict-affected region.

Vaccines delivered through this mechanism included routine antigens such as BCG, pentavalent vaccine (DTP-HepB-Hib), measles-containing vaccine, rotavirus vaccine, pneumococcal conjugate vaccine (PCV), bivalent oral polio vaccine (bOPV) and inactivated polio vaccine (IPV), as well as vaccines used in outbreak response campaigns including oral cholera vaccine (OCV) and novel oral polio vaccine type 2 (nOPV2). Table 2 presents the distribution of vaccines by state and the number of localities reached across the Darfur region.

**Table 2:**
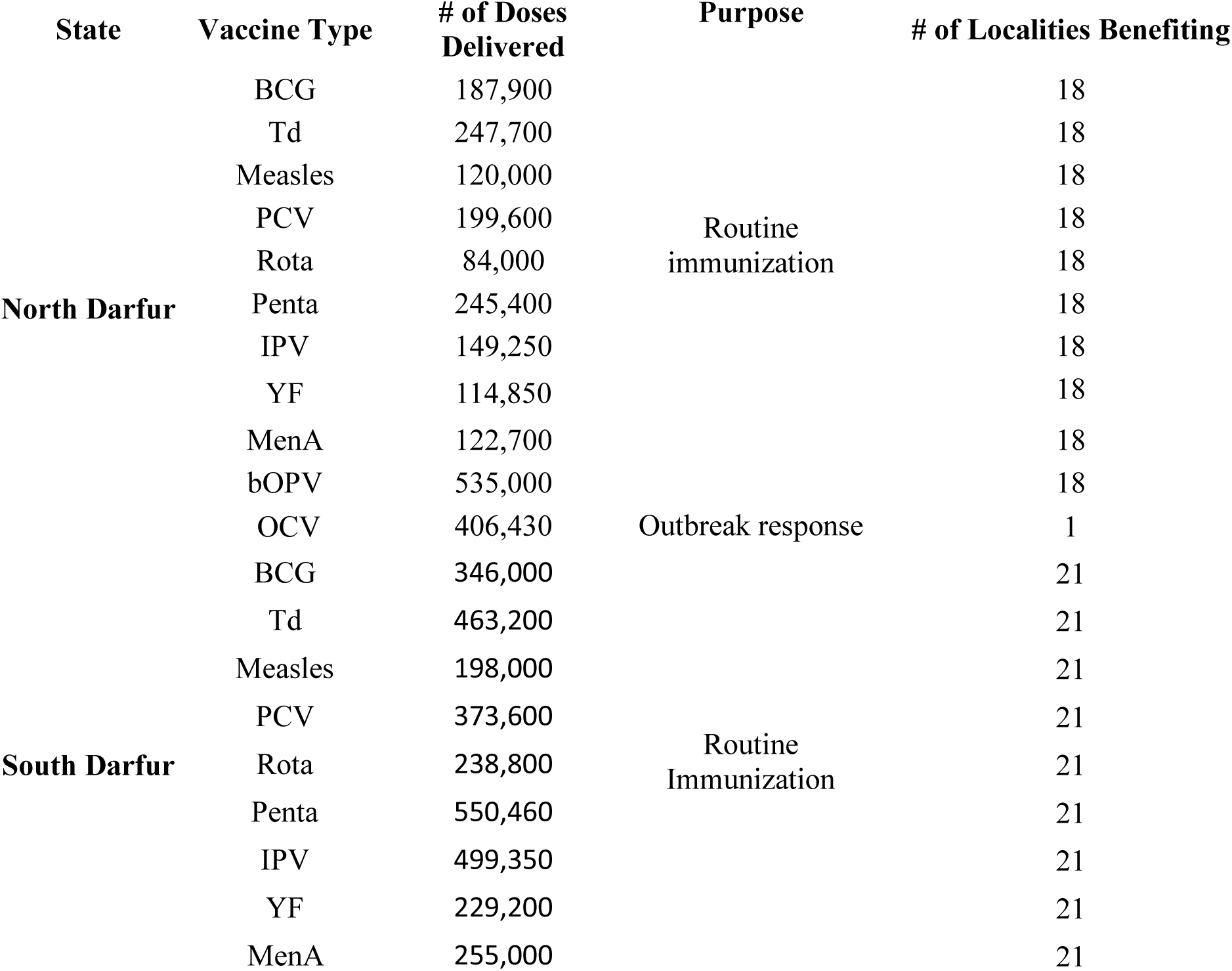
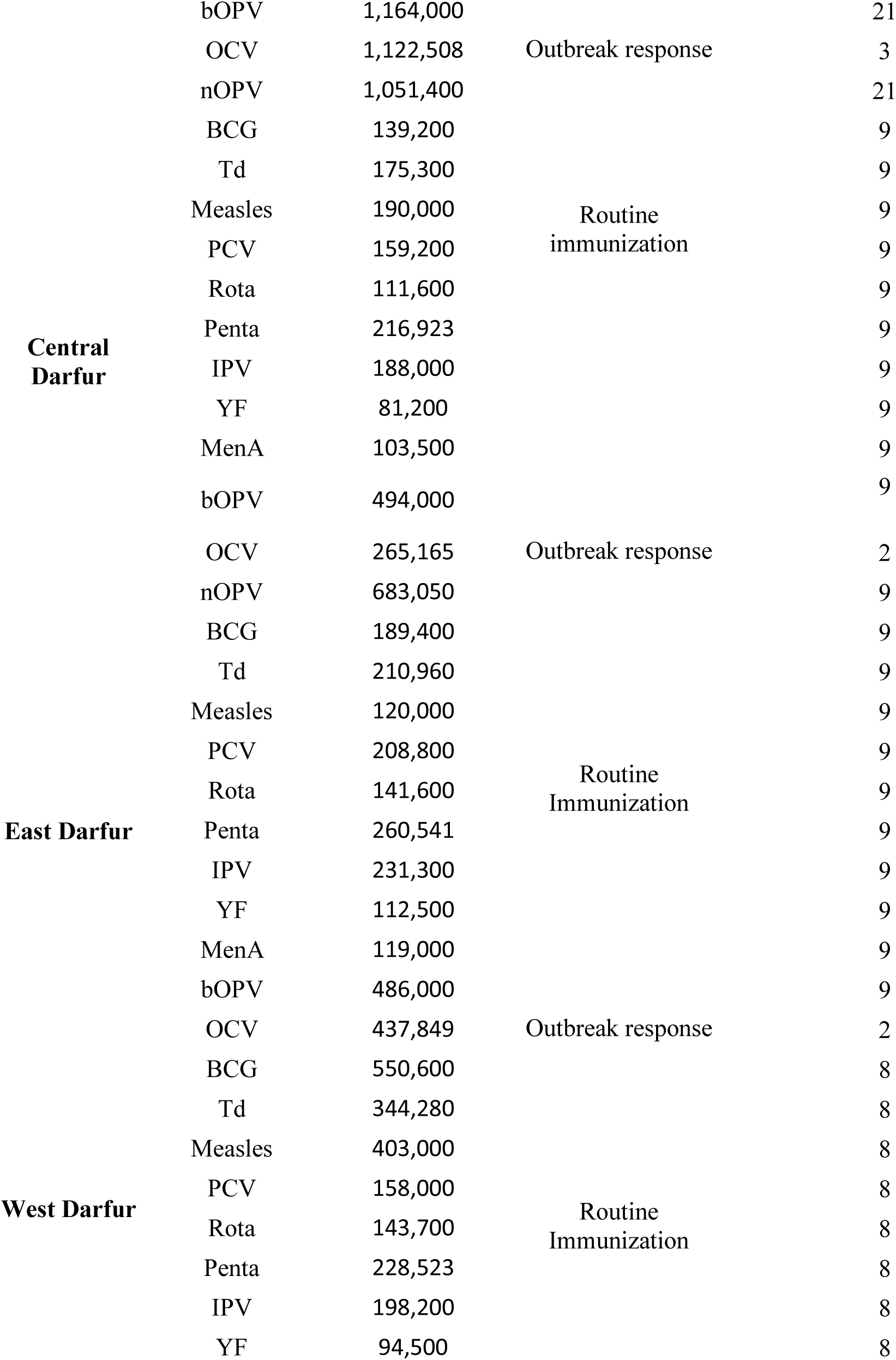
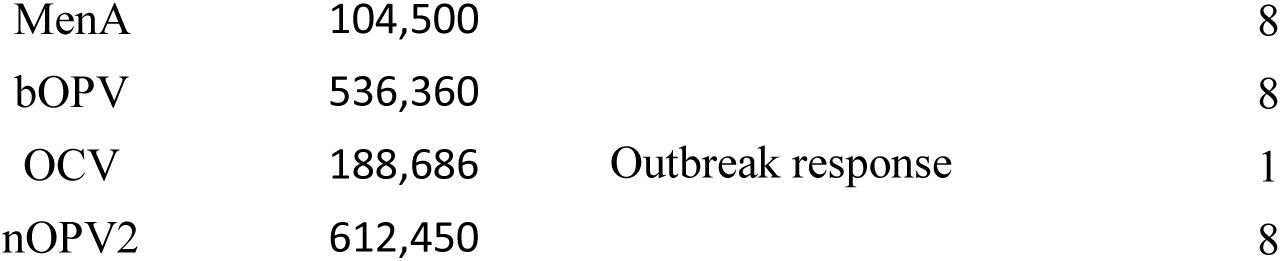
Vaccine Distribution by State and Localities covered in 2025 State.

**Table 3:** January-December 2025 Routine immunization Coverage by State.

| State | No. of Localities Implemented | Target Population (Under 1 yrs) | DPT1(%) | DPT3 (%) | MCV1 (%) |
| --- | --- | --- | --- | --- | --- |
| North Darfur | 18 | 145,216 | 64 | 41 | 27 |
| South Darfur | 21 | 196,362 | 79 | 40 | 45 |
| Central Darfur | 9 | 63,035 | 102 | 68 | 65 |
| East Darfur | 9 | 71,552 | 111 | 104 | 79 |
| West Darfur | 8 | 63,239 | 90 | 77 | 73 |
| **Total** | **65** | **539,404** | **83.2** | **55.4** | **50.4** |

**Table 4:** 2025 Polio outbreak Vaccination Campaign Results by State.

| State/Round | No. of Localities Implemented | Target Population (Under 5 yrs) | No. Vaccinated | Coverage (%) |
| --- | --- | --- | --- | --- |
| Central Darfur/R1 | 9 | 256,375 | 290,045 | 113 |
| West Darfur/R1 | 8 | 237,294 | 252,689 | 106 |
| Central Darfur/R2 | 9 | 290,045 | 333,288 | 115 |
| West Darfur/R2 | 8 | 252,689 | 291,217 | 115 |
| **Total/R1** | **17** | **493,669** | **542,734** | **110** |
| **Total/R2** | **17** | **542,734** | **624,505** | **115** |

**Table 5:** 2025 OCV Vaccination Campaign Results by State.

| **State** | **No. of Localities Implemented** | **Target Population (Under 5 yrs)** | **No. Vaccinated** | **Coverage (%)** |
| --- | --- | --- | --- | --- |
| North Darfur | 1 | 406,430 | 303,430 | 74.7 |
| South Darfur | 4 | 1,223,909 | 1,114,510 | 90.0 |
| Central Darfur | 2 | 265,165 | 265,542 | 99.9 |
| East Darfur | 2 | 437,849 | 423,873 | 96.8 |
| West Darfur | 1 | 177,739 | 163,609 | 92.1 |
| **Total** | **10** | **2,511,092** | **2,270,964** | **90.4** |

### Routine Immunization Coverage

Restoration of vaccine availability through the cross-border supply mechanism was associated with substantial improvements in routine immunization coverage across the Darfur states in 2025. Regional coverage for the first dose of a DPT-containing vaccine (DPT1), an indicator of access to immunization services, reached 83.2% compared to substantially lower levels in 2024. . Coverage varied by state, ranging from 64% in North Darfur to 111% in East Darfur.

Coverage of the third dose of a DPT-containing vaccine (DPT3) reached 55.4%, ranging from 41% in North Darfur to 104% in East Darfur. Similarly, coverage of the first dose of measles-containing vaccine (MCV1) reached 50.4%, with state-level coverage ranging from 27% in North Darfur to 79% in East Darfur. Coverage levels exceeding 100% in some states likely reflect population movement and challenges in estimating target populations in conflict-affected settings characterized by large-scale displacement.

These findings suggest that restoring vaccine availability played a critical role in enabling recovery of immunization service delivery, even in a context of ongoing insecurity and displacement.

### Outbreak Response Vaccination

In addition to supporting routine immunization services, the cross-border vaccine supply enabled the implementation of several large-scale outbreak response vaccination campaigns in the Darfur region.

### Polio Vaccination Campaigns

Two rounds of polio outbreak response campaigns were conducted in West and Central Darfur during 2025. Each round reached more than 500,000 children under five years of age, with administrative coverage exceeding 100% of the estimated target population. Coverage levels of 110% in Round 1 and 115% in Round 2 likely reflect population movements across administrative boundaries and uncertainties in population denominators.

### Oral Cholera Vaccine Campaigns

The availability of oral cholera vaccines through the cross-border supply mechanism also enabled a large-scale vaccination campaign targeting populations at risk of cholera outbreaks. Across 10 localities in the five Darfur states, the campaign reached approximately 2.27 million individuals, representing 90.4% coverage of the targeted population. Coverage varied by state but exceeded 90% in most locations, demonstrating effective campaign implementation despite challenging operational conditions.

### System-level effects

#### Improved coordination and partnership

The cross-border vaccine deployment significantly strengthened coordination and collaboration among key stakeholders, including the Ministries of Health of Sudan and Chad, UNICEF Chad, Sudan and Supply Division offices), and Gavi. A weekly coordination meeting was established for planning and implementation of shipments. This joint effort fostered a more integrated approach to immunization supply chain management, enabling timely information sharing, joint problem-solving, and harmonized decision-making. It also laid the groundwork for a more resilient and responsive cross-border public health collaboration, with improved trust and operational alignment among partners.

#### Restored Community Confidence in Immunization

The cross-border vaccine deployment played a critical role in restoring communities’ trust in immunization services across the Darfur states. By ensuring the availability of vaccines for both routine immunization and outbreak response, it enabled the resumption of demand generation activities and community engagement efforts.

Amid conflict, economic hardship, and widespread insecurity—including lack of shelter and food, communities began to recognize the life-saving value of vaccination. The urgent, unquantifiable need to protect children from vaccine-preventable diseases, combined with clear messaging on the efficacy, safety, and cost-effectiveness of vaccines, made immunization a top priority for affected populations. This renewed confidence has not only improved uptake but also reinforced the perception of immunization as an essential service, even in the most challenging humanitarian settings.

#### Challenges and Operational Constraints

Despite the overall success of the cross-border vaccine deployment, implementation faced several operational challenges related to logistics, security, governance, and health system capacity.

##### Border Access and Regulatory constraints

Delays in obtaining import and export permits, particularly within Chad, occasionally slowed shipment timelines. To address these challenges, regular coordination meetings were established between UNICEF Sudan, UNICEF Chad, and UNICEF Supply Division to monitor shipment progress and resolve administrative bottlenecks. High-level advocacy efforts by Gavi, UNICEF leadership, and Sudan’s Federal Ministry of Health were also undertaken to facilitate timely approval of cross-border shipments.

##### Security Risks Along Transport Routes

Insecurity along transport routes required continuous adaptation of logistics planning and route selection based on real-time security assessments,

##### Fragmented Governance Structures

The presence of multiple armed actors and fragmented governance structures in parts of Darfur required careful coordination and adherence to humanitarian principles to ensure safe delivery.

##### Cold Chain and Infrastructure limitations

Weak infrastructure was addressed through deployment of additional cold chain equipment and development of tailored distribution plans

##### Human Resource and Logistics Capacity Gaps

The conflict also reduced the availability of trained logistics personnel and reliable transport providers in the region. Shortages of trained personnel were mitigated through deployment of technical staff and engagement of logistics providers.

##### Cold Chain Monitoring in Insecure Areas

Monitoring cold chain performance at the last mile was particularly challenging in areas with limited access and communication infrastructure. Hence, simplified tools such as VVMs and remote supervision mechanisms were used to mitigate this challenge.

## Discussions

The cross-border deployment of vaccines to Darfur during a period of armed conflict and humanitarian crisis represents an important operational example of how immunization supply chains can be maintained in highly constrained environments. The findings demonstrate that alternative supply mechanisms can restore vaccine availability and support the continuation of both routine immunization services and outbreak response activities when national systems are disrupted. It demonstrated the power of coordinated action, innovation, and adaptability in overcoming systemic challenges related to access, security, logistics, and governance.

A key insight from this study is that immunization service disruption in conflict settings is driven not only by insecurity but also by vaccine unavailability resulting from supply chain breakdown. The substantial improvement in coverage observed in Darfur following restoration of vaccine supply suggests that availability of vaccines is a critical determinant of service delivery, even in contexts characterized by ongoing insecurity and population displacement.

Cross-border operations are critical in delivering life-saving interventions during conflict, especially when national systems are disrupted, access to affected areas is blocked, or health infrastructure is severely compromised. These operations allow humanitarian agencies to bypass front-line barriers and deliver essential services such as vaccines, medical supplies, and nutrition interventions to populations trapped by conflict or political fragmentation. When areas within a country are under the control of non-state actors or are unsafe for humanitarian access due to active fighting, cross-border operations provide a neutral and often the only feasible corridor to reach civilians. [11]

Experiences from other conflict settings highlight the importance of such approaches. In Syria, cross-border delivery of vaccines and health supplies enabled routine immunization and large-scale polio vaccination campaigns in opposition-controlled areas [11]. During the 2013–2018 conflict in South Sudan, vaccines and medical supplies were delivered through Uganda and Kenya using cross-border logistics. Insecurity in central and southern Somalia limited access, prompting humanitarian partners to implement synchronized cross-border polio vaccination campaigns with Kenya and Ethiopia to protect nomadic and mobile populations [12]. From October 2023 onward, UN agencies and partners have used El Arish Airport in Egypt as a logistical staging hub, where hundreds of tonnes of emergency aid including trauma kits, medicines, fortified biscuits, and hygiene supplies are flown in and then transported across the border into Gaza under coordinated inspection procedures[13].These experiences highlight the importance of operational flexibility and regional collaboration in maintaining essential health services during conflict.

The experience in Darfur adds to this body of evidence by demonstrating how cross-border vaccine supply can help sustain both routine immunization services and outbreak response in a protracted conflict setting. In 2025, the delivery of nearly 20 million doses of various vaccines and the marked improvement in coverage indicators illustrate the potential of such approaches to mitigate the effects of health system disruption.

The impact of improved vaccine availability is reflected in immunization coverage outcomes. Coverage for the first dose of a DPT-containing vaccine (DPT1), a key indicator of access to immunization services, averaged 83% across the five Darfur states in 2025 [14], compared to just 22.6% in 2024 [15]. This substantial improvement suggests that insecurity alone does not fully explain poor access to immunization services; rather, vaccine unavailability is a major contributing factor to service disruption in conflict settings. Similarly, coverage for the third dose of a DPT-containing vaccine (DPT3) and the first dose of measles-containing vaccine (MCV1) reached 55.4% and 50.4%, respectively, in 2025 [14], compared to 11.1% and 14.5% in 2024 [8].Although coverage levels remain below pre-conflict levels, the magnitude of improvement suggests that vaccine unavailability, in addition to insecurity, was a major driver of service disruption in the region. . These findings highlight an important insight for immunization programmes operating in conflict settings: restoring vaccine availability can significantly contribute to the recovery of service delivery even in environments where insecurity and displacement persist. Similar improvements have been reported in other conflict-affected settings, including Yemen, where enhanced humanitarian support was associated with substantial gains in immunization coverage. [15].

Cross-border deployment also enabled the implementation of oral cholera vaccine (OCV) campaigns in response to outbreaks in ten localities across the five Darfur states, achieving 90.4% coverage among the targeted population aged one year and above (2,511,092 individuals) [16]. In addition, two rounds of polio outbreak response campaigns were conducted in West and Central Darfur, reaching coverage levels exceeding 100% (110% and 115%) among children under five years of age [17]. These coverage levels highlight challenges in target population estimation, a common issue in conflict-affected settings characterized by large-scale population displacement. Comparable patterns were observed in southern Yemen, where polio campaign coverage reached 104% and 105% during two rounds conducted in 2025 [18].

Amid conflict, economic hardship, and widespread insecurity including lack of shelter and food, communities began to recognize the life-saving value of vaccination. The urgent, unquantifiable need to protect children from vaccine-preventable diseases, combined with clear messaging on the efficacy, safety, and cost-effectiveness of vaccines, made immunization a top priority for affected populations. This renewed confidence has not only improved uptake but also reinforced the perception of immunization as an essential service, even in the most challenging humanitarian settings

The intervention contributed to strengthened coordination among national authorities, international agencies, and implementing partners, with regular coordination mechanisms enabling effective information sharing, alignment of operational decisions, and timely resolution of logistical challenges. The phased and evidence-driven approach adopted for the test shipment provided a valuable learning platform, informing the design and refinement of the broader cross-border vaccine deployment strategy. These experiences highlighted the critical importance of regional collaboration, contingency planning, and adaptability in advancing immunization equity in fragile and conflict-affected settings. Overall, the findings underscore the central role of strong partnerships and operational flexibility in effectively responding to public health emergencies in complex humanitarian contexts.

There are several limitations to this study. First, the analysis is based on routinely collected programmatic data, which may be affected by inaccuracies in population denominators, particularly in settings with large-scale displacement. Second, improvements in immunization coverage observed during the study period cannot be attributed solely to the cross-border vaccine deployment, as other concurrent interventions including outreach activities, campaign-based vaccination, and partner support may also have contributed to service recovery. Finally, limited access to some areas of Darfur restricted the ability to independently verify service delivery and cold chain performance at the facility level.

Despite these limitations, the findings have important implications for immunization programs in fragile and conflict-affected settings. While cross-border operations are essential during acute emergencies especially in contexts where conflict, displacement, or state collapse has disrupted internal systems they are inherently temporary solutions. These operations serve as life-saving stopgaps when national channels fail to deliver essential services, such as vaccines, food, or medical care. However, prolonged reliance on cross-border mechanisms can undermine national ownership and weaken efforts to rebuild or strengthen in-country systems. Therefore, it is critical to view cross-border operations as part of a phased humanitarian-to-development continuum, not a parallel or substitute system. Strengthening national health systems remains essential for achieving sustainable immunization coverage. As security conditions improve, cross-border interventions should gradually transition toward nationally managed supply chains, with continued investments in infrastructure, workforce capacity, and governance mechanisms. Integrating cross-border strategies into broader preparedness planning may help ensure more resilient immunization systems capable of responding to future crises.

### Recommendations

Considering the findings and challenges highlighted, this section outlines key recommendations aimed at strengthening the effectiveness, sustainability, and coordination of cross-border health operations in fragile and conflict-affected settings.

1. **Establish Clear Regional and National Policy Frameworks:** Governments, with support from partners, should adopt or formalize policies that allow for pre-authorized cross-border humanitarian access during crises. These frameworks should define legal mandates, vaccine import protocols, and responsibilities among partners to avoid delays during emergencies. Regional agreements (e.g., IGAD or AU-level) can help standardize these across borders.
2. **Integrate Cross-Border Preparedness into National Health Emergency Plans**: Countries should incorporate cross-border scenarios in national preparedness and response plans (e.g., cholera, measles, polio). This includes mapping border communities, designing logistical corridors, and pre-positioning supplies. Scenario-based simulations and joint contingency planning with neighboring countries should be conducted regularly.
3. **Strengthen Multi-Level Coordination Mechanisms:** Effective cross-border operations require coordination at three levels: (1) international among UN agencies, donors (Gavi, WHO, UNICEF and others); (2) national between ministries of health, border control, and customs; and (3) local across implementing partners and health districts. A standing cross-border coordination task force can help maintain real-time communication, avoid duplication, and ensure joint monitoring.
4. **Promote Government Engagement and Ownership:** Early and sustained engagement with national authorities is essential. Cross-border operations should align with national strategies to avoid parallel systems and support long-term system strengthening.
5. **Strengthen Security Risk Assessments and Humanitarian Access Negotiations**: real-time security assessments and engagement with relevant actors are essential. Where appropriate, negotiations with non-state actors should be pursued under humanitarian principles to ensure safe passage of health teams and supplies. Establishing neutral humanitarian corridors coordinated with all parties to conflict is essential for continuity and safety.
6. **Strengthen Logistics Resilience and Redundancy:** Supply routes used during cross-border operations should be flexible and diversified to reduce vulnerability to security disruptions. Identifying alternative routes, staging hubs, and transport providers can improve resilience during humanitarian emergencies.

## Conclusions

Cross-border health logistics should be a central pillar of any future-oriented global health security strategy.

Cross-border vaccine deployment in Darfur demonstrates how innovative supply chain strategies can sustain essential health services during conflict. By establishing an alternative logistics corridor through Chad, partners were able to restore vaccine availability and support both routine immunization services and outbreak response campaigns in a region where internal supply routes had collapsed.

The findings highlight that vaccine availability is a critical determinant of immunization service delivery in conflict settings, and that restoring supply can enable substantial recovery even in the presence of ongoing insecurity and displacement.

The experience underscores the importance of operational flexibility, strong partnerships, and adaptive logistics systems in delivering health interventions in fragile and conflict-affected settings. Cross-border supply mechanisms can provide critical lifelines for vulnerable populations when national systems are disrupted. At the same time, such approaches should be viewed as temporary solutions that complement—not replace—national health systems. Long-term improvements in immunization coverage will ultimately depend on rebuilding resilient national supply chains and strengthening local health system capacity. As conflicts, humanitarian crises, and climate-related shocks increasingly disrupt health systems worldwide, strengthening cross-border health logistics will become an important component of global health security and emergency preparedness strategies.

## Data Availability

All data produced in the present study are available upon reasonable request to the authors

## Reference

1. Osei, E., Ibrahim, M., & Kofi Amenuvegbe, G., 2019, Effective Vaccine Management: The Case of a Rural District in Ghana. Advances in preventive medicine, 2019, 5287287. 10.1155/2019/5287287

2. Pollard, A.J., Bijker, E.M., 2021, A guide to vaccinology: from basic principles to new developments. Nat Rev Immunol 21, 83–100, 10.1038/s41577-020-00479-7

3. World Health Organization. Regional Office for Africa., 2017, Mid-Level Management Course for EPI Managers: Block III: Logistics: Module 8: Vaccine management, https://apps.who.int/iris/handle/10665/260479

4. Ameen, H., Salaudeen, A.G., Musa, O. et al., Predictors of vaccine management practices among primary healthcare workers (PHCWs) in Ilorin, North Central Nigeria. Research Journal of Health Sciences, 4. 2016–148.https://www.researchgate.net/publication/305495236_Predictors_of_vaccine_management_practices_among_primary_healthcare_workers_PHCWs_in_Ilorin_North_Central_Nigeria

5. WHO., & UNICEF., 2016, WHO/UNICEF joint statement: Achieving immunization targets with the comprehensive effective vaccine management (cEVM) framework. https://apps.who.int/iris/handle/10665/254717

6. International organization for Migration (IOM)., 2025. International Organization for Migration (IOM) Displacement Tracking Matrix (DTM)

7. Sudan., 2025. WHO and UNICEF estimates of immunization coverage: 2023 revision

8. Sudan., 2024. Ministry of Health 2019-2024 Routine Immunization administrative coverage data

9. FMOH., UNICEF Sudan., 2024. Updated Immunization supplies deployment plan for Darfur states

10. World Health Organization (2022). Reaching Every Community: Delivering Immunization in Conflict Settings. https://www.who.int/publications/i/item/9789240064307

11. OCHA., 2023. UN Cross-border Operations Northwest Syria https://www.unocha.org/publications/report/syrian-arab-republic/northwest-syria-united-nations-cross-border-operations-turkiye-syria-31-july-2023-enartr

12. WHO EMRO., 2021. Horn of Africa Cross-border Health Initiative https://www.emro.who.int/polio-eradication/news/horn-of-africa-health-ministers-commit-to-bolster-polio-eradication-efforts.html#:∼:text=Country%2Dlevel%20vaccination%20efforts,child%20and%20in%20environmental%20samples

13. UN. 2023. https://news.un.org/en/story/2023/10/1142407?utm_source=chatgpt.com

14. Sudan., 2025. Ministry of Health 2025 Routine Immunization administrative coverage data

15. Yemen., 2025. Ministry of Health 2025 Routine Immunization administrative coverage data

16. Sudan., 2025. Ministry of Health 2025 Darfur OCV campaign administrative coverage data

17. Sudan., 2025. Ministry of Health 2025 Darfur Polio campaign administrative coverage data

18. Yemen., 2025. Ministry of Health 2025 South Polio campaign administrative coverage data

